# Pharmacogenetic Study of Antipsychotic-Induced Lipid and BMI Changes in Chinese Schizophrenia Patients: A Genome-Wide Association Study

**DOI:** 10.1101/2024.09.04.24313052

**Authors:** Kenneth Chi-Yin Wong, Perry Bok-Man Leung, Benedict Ka-Wa Lee, Zoe Zi-Yu Zheng, Emily Man-Wah Tsang, Meng-Hui Liu, Kelly Wing-Kwan Lee, Shi-Tao Rao, Pak-Chung Sham, Simon Sai-Yu Lui, Hon-Cheong So

## Abstract

Second-generation antipsychotics (SGAs) are widely used to treat schizophrenia (SCZ), but they often induce metabolic side effects, including dyslipidemia and obesity, posing significant clinical challenges. While genetic factors are believed to contribute to the variability of these side effects, pharmacogenetic studies remain limited. This study aimed to identify genetic variants associated with SGA-induced lipid and BMI changes in a Chinese SCZ cohort using genome-wide association studies (GWASs). A naturalistic longitudinal cohort of Chinese SCZ patients receiving SGAs was followed for up to 18.7 years. We analyzed the patients’ genotypes (*N*=669), lipid profiles and BMI, utilizing 19 316 prescription records and 3 917 to 7 596 metabolic measurements per outcome. Linear mixed models were used to estimate the random effects of SGAs on lipid profiles and BMI changes for each patient. GWAS and gene set analyses were conducted with false discovery rate (FDR) correction. Two genome-wide significant SNPs were identified under an additive genetic model: rs6532055 in *ABCG2* (olanzapine-induced LDL changes) and rs2644520 near *SORCS1* (aripiprazole-induced triglyceride changes). Three additional SNPs achieved genome-wide significance under non-additive models: rs115843863 near *UPP2* (clozapine-induced HDL changes), rs2514895 near *KIRREL3* (paliperidone-induced LDL changes), and rs188405603 in *SLC2A9* (quetiapine-induced triglyceride changes). Gene-based analysis revealed six genome-wide significant (p<2.73e-06, Bonferroni correction) genes: *ABCG2*, *APOA5*, *ZPR1*, *GCNT4*, *MAST2*, and *CRTAC1*. Four gene sets were significantly associated with SGA-induced metabolic side effects. This pharmacogenetic GWAS identified several genetic variants associated with metabolic side effects of seven SGAs, potentially informing personalized treatment strategies to minimize metabolic risk in SCZ patients.

## 1. Introduction

Schizophrenia (SCZ) is a severe, chronic mental illness with high heritability and a lifetime prevalence of approximately 1%. The global burden of SCZ has been increasing, with the incidence increasing by 2% annually between 2000 and 2019^1–3^. Cardiovascular disease (CVD) is the leading cause of mortality in SCZ patients^4^, and psychosis itself is also a recognized risk factor for dyslipidemia and obesity^5^. Moreover, second-generation antipsychotics (SGAs), the mainstream treatment for SCZ, can adversely affect patients’ lipid profiles, other metabolic parameters, and body mass index (BMI)^6^. Interestingly, the propensity to develop these metabolic side effects varies considerably among individuals. Twin and sibling studies have demonstrated that such interindividual variability may be largely attributable to genetic differences^7, 8^. However, the underlying genetic mechanisms remain poorly understood.

Pharmacogenetics (PGx) examines how genetic variations affect drug metabolism and response, potentially enabling personalized treatment plans with better efficacy and fewer side effects. Over the past two decades, most PGx studies on SGA-induced metabolic side effects have employed candidate gene approaches, focusing primarily on dopamine and serotonin receptor-related genes^9, 10^. Additionally, variants in cytochrome P450 genes (*CYP1A2*, *CYP2C19*, *CYP2D6* and *CYP3A4*) have been reported to be associated with antipsychotic serum concentrations^11, 12^. The advent of genome-wide association studies (GWASs) has largely overcome the limitations of candidate gene approaches, uncovering more variants and genes associated with antipsychotic response^13–17^. However, the majority of previous PGx studies have focused mainly on treatment response rather than the metabolic side effects of SGAs.

To date, only seven PGx GWASs have been conducted to investigate SGA-induced metabolic side effects^18–24^. All but one study focused exclusively on weight gain, with Adkins *et al*. (2010) investigating PGx on a variety of metabolic side effects across five SGAs and one typical antipsychotic^18^. Additionally, most studies focused on short-term outcomes, with the longest follow-up reported by Adkins *et al*. (18 months) ^18^. Nevertheless, the work of Adkins *et al.* has several limitations, as acknowledged by the original authors^18^. For example, most participants had prior experience with antipsychotics, and many were concurrently taking other medications, such as antidepressants and mood stabilizers, which could potentially influence the results. Notably, the subjects had received, on average, 14.3 years of antipsychotic medications at baseline. Moreover, DNA collection was conducted after the clinical trial, involving only a subset of participants (51%) who exhibited lower symptom severity and reduced rates of drug and alcohol dependence, potentially introducing bias. With respect to GWAS analysis, Adkins *et al*. did not employ imputation, which may limit the ability to detect true genetic associations.

To address this knowledge gap, our PGx study investigated lipid and BMI changes induced by seven SGAs: olanzapine (OZP), clozapine (CZP), quetiapine (QUE), risperidone (RIS), aripiprazole (ARI), amisulpride (AMI) and paliperidone (PAL). We focused on BMI and four lipid measurements, including total cholesterol (TC), high-density cholesterol (HDL), low-density cholesterol (LDL) and triglycerides (TG), as outcomes. Compared with Adkins *et al*.’s previous study which followed patients for 1.5 years, we utilized a longitudinal cohort with a longer follow-up of up to 5.7 years (median), and our naturalistic dataset had a greater mean number of metabolic measures per subject. The proportion of SGA-naïve patients was also markedly greater at ∼63%. Furthermore, the homogeneity of our Chinese cohort, which was recruited from Hong Kong, China, combined with imputed genotypes based on the ChinaMAP reference panel, enhanced the statistical power of GWAS to detect true signals.

Taken together, this sophisticated approach combined with a long follow-up duration and comprehensive medication history and metabolic measures. We aimed to identify novel genetic variants associated with SGA-induced metabolic side effects. Our findings provide insights into the biological mechanism underlying SGA-induced lipid and BMI changes, potentially contributing to more personalized and effective treatments for SCZ patients.

## 2. Subjects and Methods

### 2.1 Study population and data collection

We recruited SCZ patients from an early psychosis intervention clinic at Castle Peak Hospital Hong Kong between 2009 and 2021^25^. The inclusion criteria were as follows: (1) aged ≥18 years, (2) Chinese ethnicity, (3) ICD-10 diagnosis of SCZ or schizoaffective disorder, (4) treatment with SGAs, and (5) at least one post-SGA measurement of fasting lipids and/or BMI. We excluded patients with preexisting metabolic disorders or those lost to psychiatric follow-up as of March 2021.

From 767 eligible patients, we extracted complete medication records, lipid profiles and BMI measurements from initial service contact to the study endpoint. Following local guidelines for monitoring SGA side effects, patients received baseline measures of fasting lipid profiles and BMI before SGA initiation, with annual follow-up measurements while on SGAs. Electronic health records documented the type and dosage of all psychotropic and concomitant medications, including antidepressants and lipid-lowering drugs.

### 2.2 Genotyping and imputation

Blood samples from patients were genotyped using the Illumina Asian Screening Array-24 v1.0. Quality control was performed using PLINK 1.9p, removing genotypes and subjects based on missing data (missing rate > 10%), Hardy‒Weinberg equilibrium (p < 1e-06), and relatedness (IBS distance > 0.25). No ethnic outliers were identified through principal component analysis.

The genome coordinates were lifted from GRCh37 to GRCh38 using CrossMap v0.6.4. Haplotype phasing and genotype imputation were conducted using Eagle2 and Minimac4, respectively^26, 27^, with the ChinaMAP phase 1.v1 reference panel (59 010 860 sites from 10 155 Chinese individuals)^28^. This large, ancestrally matched reference panel improved imputation accuracy. The imputed SNPs were removed based on imputation quality (INFO score ≤ 0.3) and minor allele count (MAC ≤ 10). The final dataset comprised 6 992 805 high-quality imputed SNPs.

### 2.3 Data preprocessing and variable selection

Full GWAS data were available for 669 SCZ patients with 19 316 prescription records within 3 months of metabolic measurement. We analyzed TC, HDL, LDL, and TG (all in mmol/L), and BMI (kg/m^2^) separately. Outliers exceeding six standard deviations from the group mean were removed to avoid a biased effect estimation. As a result, the final analysis included 4 048 TC, 3 917 HDL, 4 035 LDL, 4 034 TG, and 7 596 BMI records. We applied natural log transformation to the data prior to modeling.

We selected seven SGAs that had been prescribed to at least 30 patients in our sample, namely CZP, OZP, ARI, AMI, PAL, RIS, and QUE (**Table 1**). The choices of these SGAs, which were taken by a substantial number of subjects, allowed more robust models to be constructed. Long-acting injectable and oral formulations were analyzed equally, after dose conversion using the standard method^29^. Given that some non-SGA psychotropics and statins (lipid-lowering drugs) are commonly prescribed, and might influence patients’ lipid profiles and BMI, we accounted for these concomitant medications (i.e., haloperidol, valproate, lithium, metformin, simvastatin, and atorvastatin) by including them as covariates in our model, as suggested by prior studies^30–33^. We also included daily drug dosage (mg) and treatment duration (month) as random-effect covariates, while age, gender and years of education were entered as fixed-effect covariates.

**Table 1.** Descriptive statistics of the second-generation antipsychotic prescription records in our cohort.

| Second Generation<br>Antipsychotics (SGAs)<sup>[1]</sup> | No. of subject with prescription |  | Total no. of prescription |  | Avg. no. of prescription per subject (SD) |  |
| --- | --- | --- | --- | --- | --- | --- |
|  | TC,LDL,HDL,TG<br>(N=625) | BMI (N=646) | TC,LDL,HDL,TG<br>(N=3525) | BMI<br>(N=6640) | TC,LDL,HDL,TG<br>(N=740) | BMI (N=759) |
| **CLOZAPINE** | 119 | 125 | 551 | 1,045 | 4.6 (2.8) | 8.4 (10.1) |
| **OLANZAPINE** | 230 | 307 | 760 | 1,447 | 3.3 (2.6) | 4.7 (4.5) |
| **ARIPIPRAZOLE** | 238 | 275 | 728 | 1,228 | 3.1 (2.2) | 4.5 (4.3) |
| **RISPERIDONE** | 116 | 232 | 209 | 571 | 1.8 (1.7) | 2.5 (2.5) |
| **AMISULPRIDE** | 162 | 216 | 399 | 766 | 2.5 (2.0) | 3.5 (3.2) |
| **QUETIAPINE** | 93 | 105 | 173 | 313 | 1.9 (1.2) | 3 (3.1) |
| **PALIPERIDONE** | 43 | 65 | 95 | 178 | 2.2 (1.8) | 2.7 (2.3) |
| **Any SGA use** | 553 | 567 | 2,591 | 5,259 | 4.7 (2.9) | 9.3 (8.8) |

### 2.4 Random-effect estimation for SGA-induced lipid/BMI changes

We used linear mixed models (LMMs) to estimate the random effects (random slope) of SGAs on lipid/BMI changes. The random effects quantified how each patient’s lipid/BMI changes deviated from the cohort’s mean, serving as a proxy outcome measure for the severity of the metabolic side effects of each patient. A similar approach has been employed in several PGx GWASs^18, 34, 35^. In addition, we employed an advanced statistical approach to disentangle the within-subject estimates from the between-subject estimates of SGA random effects ^36–38^. By focusing on the within-subject effects, we can more accurately estimate the metabolic side effects of SGAs by accounting for unmeasured time-invariant confounders^39^.

For each of the seven SGAs (plus one additional model of ‘any SGA use’), five random-effect LMMs were built with corresponding lipid/BMI measures as outcomes, resulting in 40 models. Random effect coefficients were extracted for patients prescribed the corresponding SGAs from these 40 models with their specifications listed in **Supplementary Text 1.** Following other PGx studies^18, 34, 35^, the random-effect coefficients were the primary outcome of the subsequent GWAS and MAGMA analyses, with sample sizes varying across models (**Table 1**). Details concerning how we identified the best-fitting random-effect LMM models and estimated the within-subject dosage of the SGAs have been described elsewhere^6, 39^.

We applied rank-based inverse normal transformation (INT) to the random-effect coefficients^40^ to ensure a normal distribution of the outcomes (and residuals) and reduce outlier effects (**Supplementary Figure S1**).

### 2.5 Genome-wide association study (GWAS) analysis

GWAS association tests between SNP dosages and SGA-induced lipid/BMI changes were conducted using PLINK 2.00a^41^, with gender and the top ten genetic principal components as covariates. The imputed genotypes were converted to PLINK 2 binary formats to retain dosage information, which can improve the statistical power of the association tests. In our primary analyses, we tested additive genetic models using allelic dosage as the predictor. To capture variants with non-additive genetic effects as advised by Guindo-Martínez, Amela ^42^, we also performed additional analyses based on dominant, recessive and genotypic (2 degrees of freedom) models.

Associations with a p-value (p) < 5e-08 were considered genome-wide (GW) significant^43^, whereas those with p ≥ 5e-08 but a false discovery rate (FDR) < 0.2 were considered ‘suggestive’ associations. A similar approach was also employed by Adkins *et al*. in their GWAS of the metabolic side effects of antipsychotics^18^.

### 2.6 MAGMA analyses

MAGMA is a powerful approach for testing the associations between a phenotype and SNPs aggregated within an entire gene^43^. We performed gene and gene-set association tests between the imputed genotypes and 40 sets of random-effect coefficients of SGA-induced lipid/BMI changes using MAGMA v1.10. To leverage the strengths and mitigate the weaknesses of each MAGMA model, we built three predefined MAGMA models, including (1) principal component regression, (2) the SNP-wise mean, and (3) the SNP-wise top 1. MAGMA then merged the resulting gene p-values into a single aggregated p-value. This approach has increased the statistical power and sensitivity across various genetic architectures^43^. Such MAGMA models have been detailed in the MAGMA manual.

### 2.7 Post-GWAS annotation

LD-clumping was performed using PLINK to identify top SNPs within linkage disequilibrium clusters (with clump-p1=5e-05, clump-p2=0.05, r2=0.6 and window size=250 kb). The top SNPs were annotated using Ensembl Variant Effect Predictor (VEP) v111.0 with VEP cache version 111_GRCh38^44^, including gene information, nearest gene, location, and effect allele frequency in East Asian and European populations. Previous studies reporting GW-significant SNPs within the same genes were annotated based on the GWAS Catalog and Open Target Platform^45, 46^. To uncover potential hidden associations between the identified genes and annotated enriched terms across multiple datasets and resources, integrative gene set enrichment analyses and visualisation were conducted using the Enrichr-KG platform, incorporating four gene-set libraries^47^: GWAS Catalog (2019), GO_biological Process (2021), DisGeNET, and Human Phenotype Ontology. For each input gene set, the top five enriched terms per library with an FDR < 0.05 were considered significant. A subnetwork linking the input genes to these enriched terms was visualized using the Enrichr-KG platform.

### 2.8 Fine-mapping with the SuSiE model

To identify potential causal SNPs among the GW-significant findings, we employed the SuSiE fine-mapping approach^48^. This method is built on top of variable selection in a linear regression model, fitted by a Bayesian analog of stepwise selection methods. This method reports as many groups of SNPs (credible sets) as the data support, each with the minimum number of SNPs possible. For each credible set, it estimates the posterior distribution of the SNPs’ effect and calculates marginal posterior inclusion probabilities (PIPs) to assess the strength of evidence for each SNP being causal. The region (±1 000 kb) around each GW-significant SNP was taken for fine-mapping using the LD reference panel constructed from the subject’s imputed genotypes. The SuSiE model fit was based on the p-value of the SNPs, with the maximum number of nonzero effects (L) set to 11, where default values were used for the remaining parameters. The casual SNPs of the best credible sets were visualized via region plots.

### 2.9 Multiple testing correction

We employed the FDR approach to balance the risk of false discoveries against potential true findings. We reported the FDR separately for each analysis, which controlled the expected proportion of false discoveries among rejected null hypotheses^49^. In contrast to Bonferroni correction, the FDR is much less affected by the number of tests, because the FDR controls the proportion (instead of the number) of false discoveries^50, 51^. Intuitively, when controlling the FDR at 0.2 for example, at least 80% of the significant findings would be “true discoveries” on average.

Since the FDR controls the proportion of false discoveries, if the FDR is performed separately for each set of analyses, the *overall* FDR is still generally controlled, especially when the number of hypothesis tests is large, as shown by Efron ^51^.

### 2.10 Ethics statement

This study adhered to the ethical principles of the Helsinki Declaration and relevant national and institutional guidelines for human research. Ethical approval was obtained from the New Territories West Cluster Ethics Committee (Approval Numbers: NTWC/CREC/823/10 and NTWC/CREC/1293/14) and the Joint Chinese University of Hong Kong-New Territories East Cluster Clinical Research Ethics Committee (Approval Number: 2016.559). All participants provided written informed consent. The software and tools used in this study are listed in **Supplementary Text 2**.

## 3. Results

### 3.1 Sample characteristics

Our final dataset comprised 625 subjects with lipid profile data and 646 subjects with BMI data after the seven SGAs prescribed to at least 30 patients were selected. **Supplementary Table S1** presents the gender ratio, mean age at the first clinical visit, and mean years of follow-up. The longitudinal cohort had a maximum follow-up period of 18.7 years, with a mean follow-ups of 5.7 years (SD = 3.3) for the lipid cohort and 5.5 years (SD = 3.2) for the BMI cohort.

We conducted 40 separate GWASs to examine the effects of SNPs on SGA-induced changes in lipids (TC, HDL, LDL, TG) and BMI for seven specific SGAs and any SGA use. The individual GWAS sample sizes ranged from 43 to 567 patients, with mean prescriptions per patient ranging from 1.8 (SD=1.7) to 9.3 (SD=8.8) (**Table 1**).

### 3.2 GWAS results

#### 3.2.1 Primary analyses: Additive genetic model

In our main GWAS analyses using an additive genetic model, two SNPs reached GW significance (p < 5e-08) (**Table 2**), whereas eight SNPs achieved an FDR of < 0.2 (Supplementary **Table S2**). The top SNP, rs6532055 (p = 3.13e-09, FDR = 0.022), was associated with olanzapine-induced LDL changes. This SNP is located within an intron of the *ABCG2* gene, which is part of the ATP-binding cassette (ABC) family. The second GW-significant SNP, rs2644520 (p = 3.06e-08, FDR = 0.122), was associated with aripiprazole-induced TG changes and located in an intergenic region near *SORCS1*, a gene encoding a member of vacuolar protein sorting 10 (VPS10) domain-containing receptor proteins.

**Table 2.**
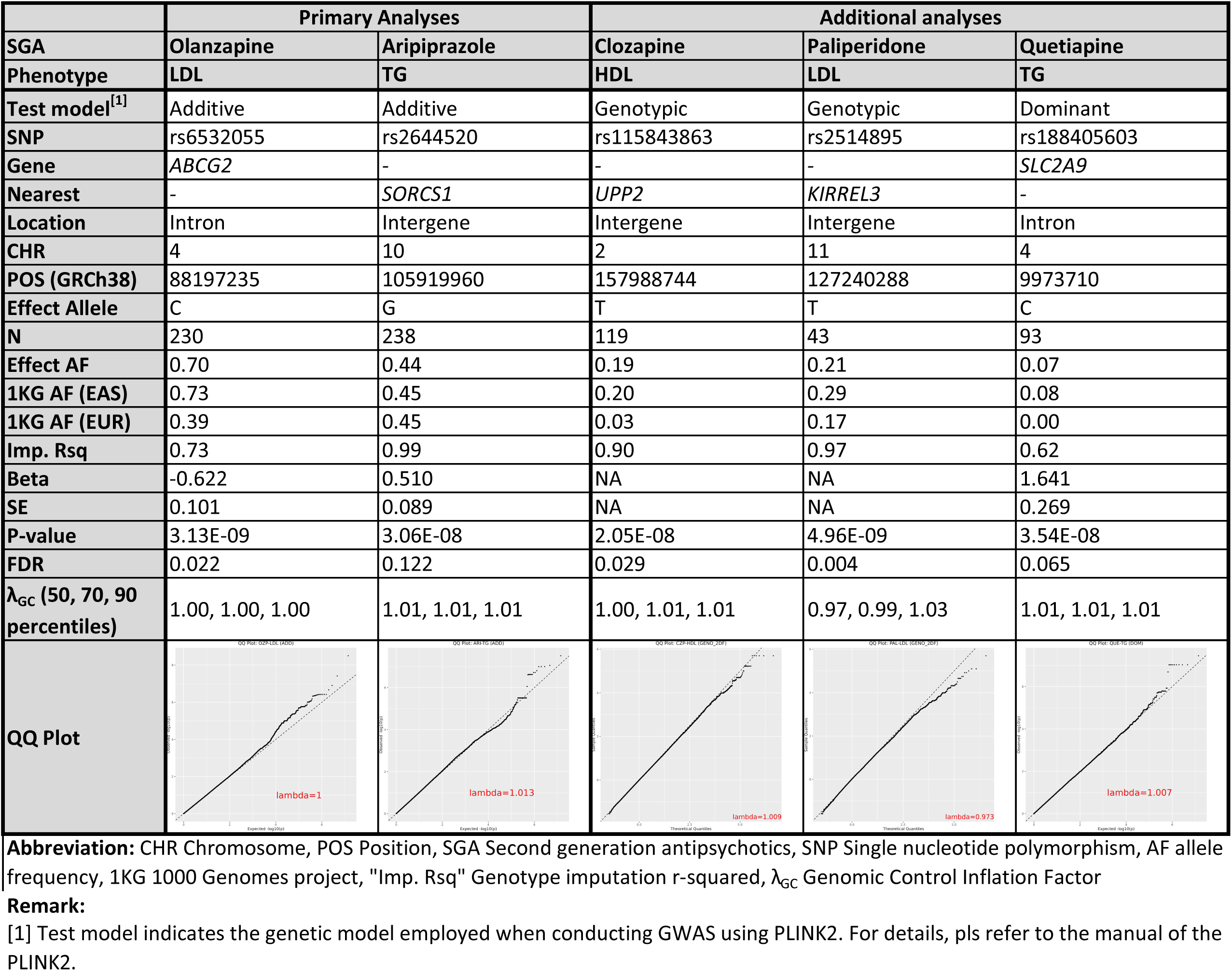
The genome-wide significant SNPs (p < 5e-08) associated with the SGA-induced lipid level and BMI changes.

|  | Primary Analyses |  | Additional analyses |  |  |
| --- | --- | --- | --- | --- | --- |
| SGA | Olanzapine | Aripiprazole | Clozapine | Paliperidone | Quetiapine |
| Phenotype | LDL | TG | HDL | LDL | TG |
| Test model<sup>[1]</sup> | Additive | Additive | Genotypic | Genotypic | Dominant |
| SNP | rs6532055 | rs2644520 | rs115843863 | rs2514895 | rs188405603 |
| Gene | ABCG2 | - | - | - | SLC2A9 |
| Nearest | - | SORCS1 | UPP2 | KIRREL3 | - |
| Location | Intron | Intergene | Intergene | Intergene | Intron |
| CHR | 4 | 10 | 2 | 11 | 4 |
| POS (GRCh38) | 88197235 | 105919960 | 157988744 | 127240288 | 9973710 |
| Effect Allele | C | G | T | T | C |
| N | 230 | 238 | 119 | 43 | 93 |
| Effect AF | 0.70 | 0.44 | 0.19 | 0.21 | 0.07 |
| 1KG AF (EAS) | 0.73 | 0.45 | 0.20 | 0.29 | 0.08 |
| 1KG AF (EUR) | 0.39 | 0.45 | 0.03 | 0.17 | 0.00 |
| Imp. Rsq | 0.73 | 0.99 | 0.90 | 0.97 | 0.62 |
| Beta | -0.622 | 0.510 | NA | NA | 1.641 |
| SE | 0.101 | 0.089 | NA | NA | 0.269 |
| P-value | 3.13E-09 | 3.06E-08 | 2.05E-08 | 4.96E-09 | 3.54E-08 |
| FDR | 0.022 | 0.122 | 0.029 | 0.004 | 0.065 |
| $\lambda_{GC}$ (50, 70, 90 percentiles) | 1.00, 1.00, 1.00 | 1.01, 1.01, 1.01 | 1.00, 1.01, 1.01 | 0.97, 0.99, 1.03 | 1.01, 1.01, 1.01 |
| QQ Plot |  |  |  |  |  |

**Abbreviation:** CHR Chromosome, POS Position, SGA Second generation antipsychotics, SNP Single nucleotide polymorphism, AF allele frequency, 1KG 1000 Genomes project, "Imp. Rsq" Genotype imputation r-squared, $\lambda_{GC}$ Genomic Control Inflation Factor
**Remark:** [1] Test model indicates the genetic model employed when conducting GWAS using PLINK2. For details, pls refer to the manual of the PLINK2.

The quantile–quantile plots (QQ) plots for the GWASs with GW-significant SNPs are shown at the bottom of **Table 2**, and the QQ plots of the GWASs with SNPs achieving an FDR < 0.2 are shown in Supplementary **Figure S2**. The QQ plots demonstrate that the p value distributions closely matched the expected p-values under the null hypothesis, with genomic control inflation factor (λ_GC_ at the median) ranging from 0.97 to 1.01 for the GWASs harboring GW-significant SNPs and ranging from 0.94 to 1.01 for the GWASs harboring SNPs with FDR < 0.2, indicating that genomic inflation is unlikely to be a concern.

#### 3.2.2 Additional analyses with non-additive models

Further analyses using dominant, recessive and genotypic models revealed three additional GW-significant SNPs (**Table 2**). The top SNP in the genotypic model, rs115843863 (p = 2.05e-08, FDR = 0.0287), was associated with clozapine-induced HDL changes. The SNP is located in an intergenic region near *UPP2*, a gene involved in dCMP and uridine catabolic processes. Another GW-significant SNP in the genotypic model, rs2514895 (p = 4.96e-09, FDR = 0.004), was associated with paliperidone-induced LDL changes and is located near *KIRREL3*, a gene encoding a nephrin-like protein expressed in the brain. The last GW-significant SNP rs188405603 (p = 3.52e-08, FDR = 0.065) under the dominant model was associated with quetiapine-induced TG changes and is located within an intron of *SLC2A9*, a gene encoding a glucose transporter.

#### 3.2.3 Suggestive associations with FDR < 0.2

Eight SNPs achieved an FDR < 0.2 in the primary GWAS analyses under an additive model (Supplementary **Table S2**). Notably, four SNPs, namely rs7412 (FDR = 0.182), rs2384157 (FDR = 0.195), rs74625905 (FDR = 0.195) and rs56349742 (FDR = 0.195), were associated with olanzapine-induced LDL changes. The well-known LDL-altering SNP rs7412 in *APOE* is positively associated with olanzapine-induced LDL changes. Four SNPs, namely rs2358259 (FDR = 0.123), rs10174314 (FDR = 0.123), rs117416034 (FDR = 0.123), and rs6424242 (FDR = 0.186), were associated with quetiapine-induced HDL changes. In particular, rs6424242 is located in an upstream region of the *SIPA1L2* gene, which has been previously linked to obesity-related traits, response to alcohol consumption, and neuroticism based on Open Targets and the GWAS Catalog^52–54^.

Another 17 SNPs with FDRs < 0.2 were identified in the additional GWAS analyses under non-additive models. These SNPs are related to the metabolic side effects of clozapine, olanzapine, risperidone, and paliperidone (**Supplementary Table S3**). The associated genes are implicated in psychiatric disorders, lipid or BMI measurements, or drug responses, including *BICD1* and *CSMD1* (olanzapine-induced TC changes); *GADL1* (risperidone-induced BMI changes); *SIPA1L2* (quetiapine-induced HDL changes); and *RAB38*, *CDH23*, *AMPH*, *FOXN3*, *APBB2*, *C1R* and *LRCOL1* (paliperidone-induced LDL changes). Additionally, GWAS analyses were conducted on the heterozygous-only (HETONLY) genetic model, revealing two GW-significant SNPs and six SNPs with suggestive evidence (FDR < 0.2). The detailed results can be found in **Supplementary Table S4**, and an overview of all identified genes from different types of analyses is provided in **Table 3**.

**Table 3.**
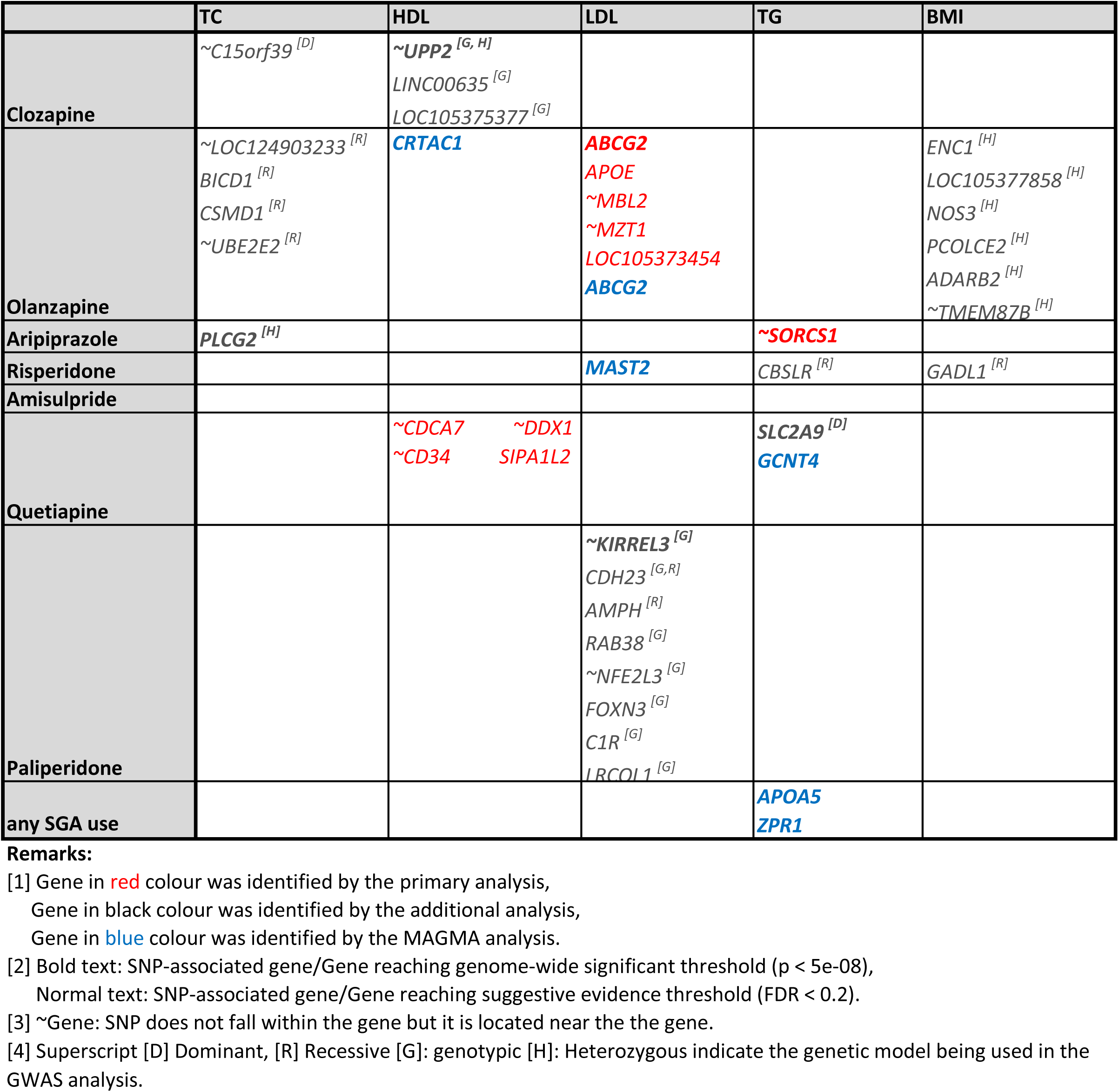
Genes associated with the SGA-induced lipid/BMI changes in different type of analyses^[1,2,3,4]^.

#### 3.2.4 Fine-mapping results

The fine-mapping results for the five GW-significant SNPs are visualized in region plots (**Figure 1**). The top GW-significant SNPs associated with olanzapine-induced LDL changes and aripiprazole-induced TG changes were proposed to be causal (PIP = 1.0). However, the remaining three top GW-significant SNPs were not considered causal, as shown by their low PIP values. Another SNP, rs73968514 (PIP = 1.0), was identified as potentially causal for clozapine-induced HDL changes (PIP = 1.0), replacing the original GWAS hit rs115843863. Both rs2441693 and another SNP rs2441693, with the same p-value, were identified as potentially causal for paliperidone-induced LDL effects (PIP = 0.5 each). Finally, instead of the observed GWAS hit rs188405603, fine-mapping evidence suggested that rs77140241 was the real causal variant for quetiapine-induced TG changes (PIP = 1.0, p = 9.5e-08, FDR = 0.065).

**Figure 1.**
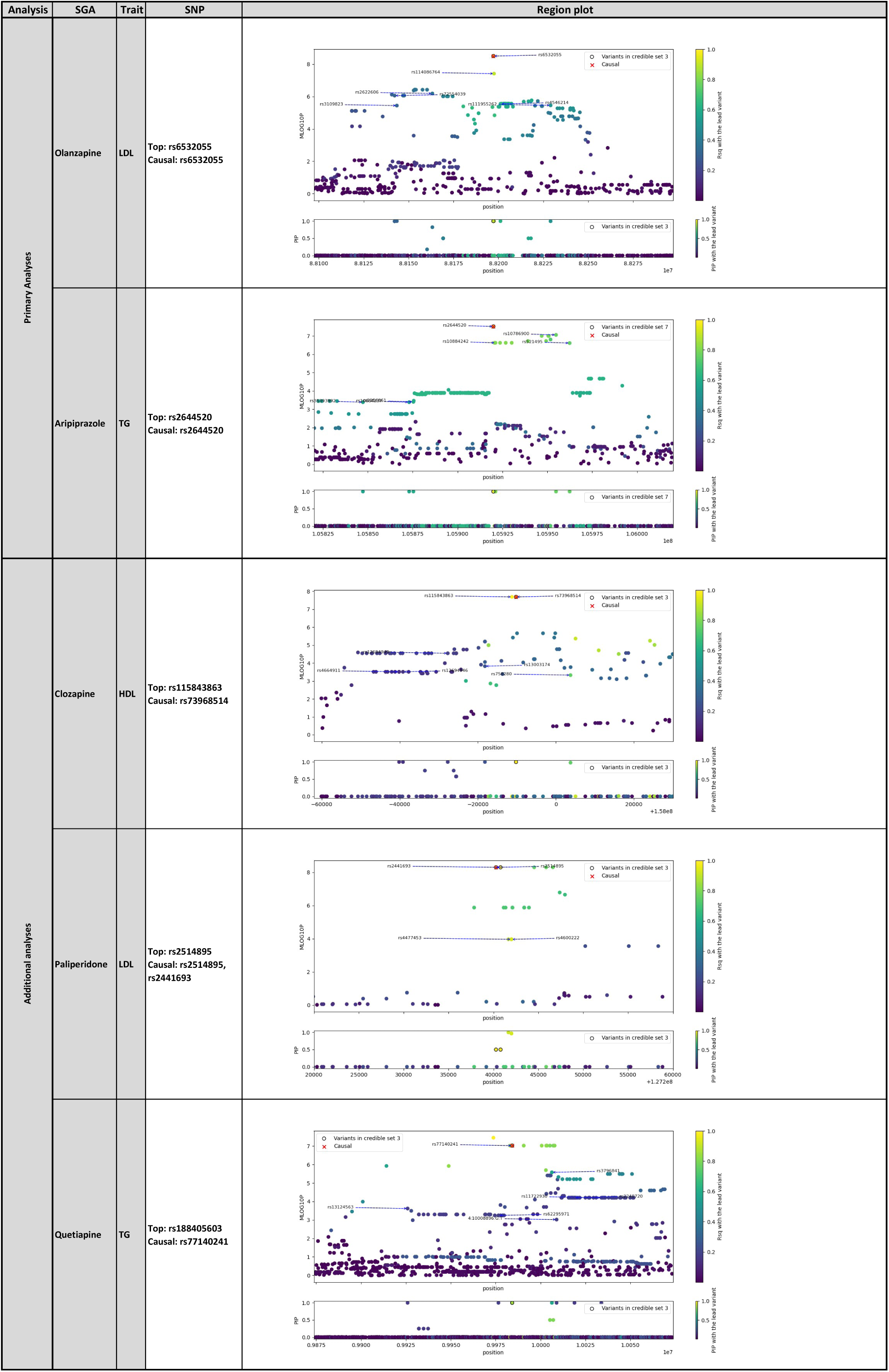
Region plots of the five GW-significant SNPs fine-mapped via the R package suiseR. The fine-mapping results of the five GW-significant SNPs are visualized in the region plots above. The lead variant, marked in yellow, is the SNP with the lowest p values after the LD-clump, whereas the causal variant is marked by a red cross. Other SNPs are coloured according to their decreasing R-square values with the lead variant. The credible set containing SNP(s) nearest to the lead variant with the smallest p values is chosen as causal SNP(s). The subplot at the bottom in each region plot shows the PIP values for the SNPs in all creditable sets, with the causal SNPs in the optimum credible set outlined by a circle.

### 3.3 MAGMA analysis results

#### 3.3.1 Gene-level analysis

Six genes reached the GW significance threshold of p < 2.73e-06 after Bonferroni correction (α = 0.05/18 288 genes tested) in the gene-level analysis(**Table 4**), with their corresponding QQ plots from the gene-level analysis shown in Supplementary **Figure S3**. All GW-significant genes also had an FDR < 0.05. Diseases or traits associated with these genes were annotated using the Open Target Platform^55^, which we also highlighted here. The top gene *ABCG2* (p = 8.26e-09, FDR = 1.51e-04) was associated with olanzapine-induced LDL changes; this gene is related to gout, urate measurement, and BMI based on information from the Open Target Platform. *APOA5* (p = 3.45e-08, FDR = 6.31e-04) and *ZPR1* (p = 1.80e-06, FDR = 0.016) were associated with SGA-induced TG changes; these genes were related to TG, HDL, and LDL levels and metabolic syndrome. *GCNT4* (p = 3.17e-07, FDR = 5.12e-03) and *MAST2* (p = 4.79e-07, FDR = 8.62e-03) were associated with quetiapine-induced TG and risperidone-induced LDL changes respectively. *CRTAC1* (p = 2.273e-06, FDR = 0.042) was associated with olanzapine-induced HDL changes. Based on the evidence from the Open Target Platform^55^, *GCNT4* and *MAST2* are related to neurodegenerative disease and measurements of erythrocyte count, BMI, LDL and TC^56–60^; whereas *CRTAC1* is related to body fat percentage and measurements of HDL and TG^60, 61^.

**Table 4.**
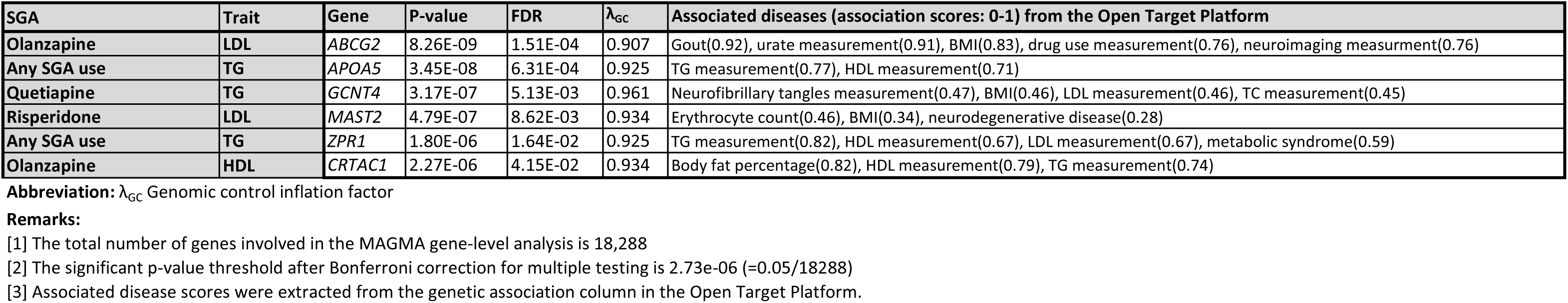
Significant gene associations with SGA-induced lipid level and BMI changes identified in MAGMA gene-level analyses (p < 2.73e-06)

Gene set enrichment analysis, incorporating these six significant genes along with those associated with the five GW-significant SNPs identified in the primary GWAS analyses, was performed using the Enrichr-KG platform. The subnetwork of gene and enriched terms are illustrated in Figure 2, with corresponding enrichment p-values and FDRs listed in Supplementary Table S5.

**Figure 2.**
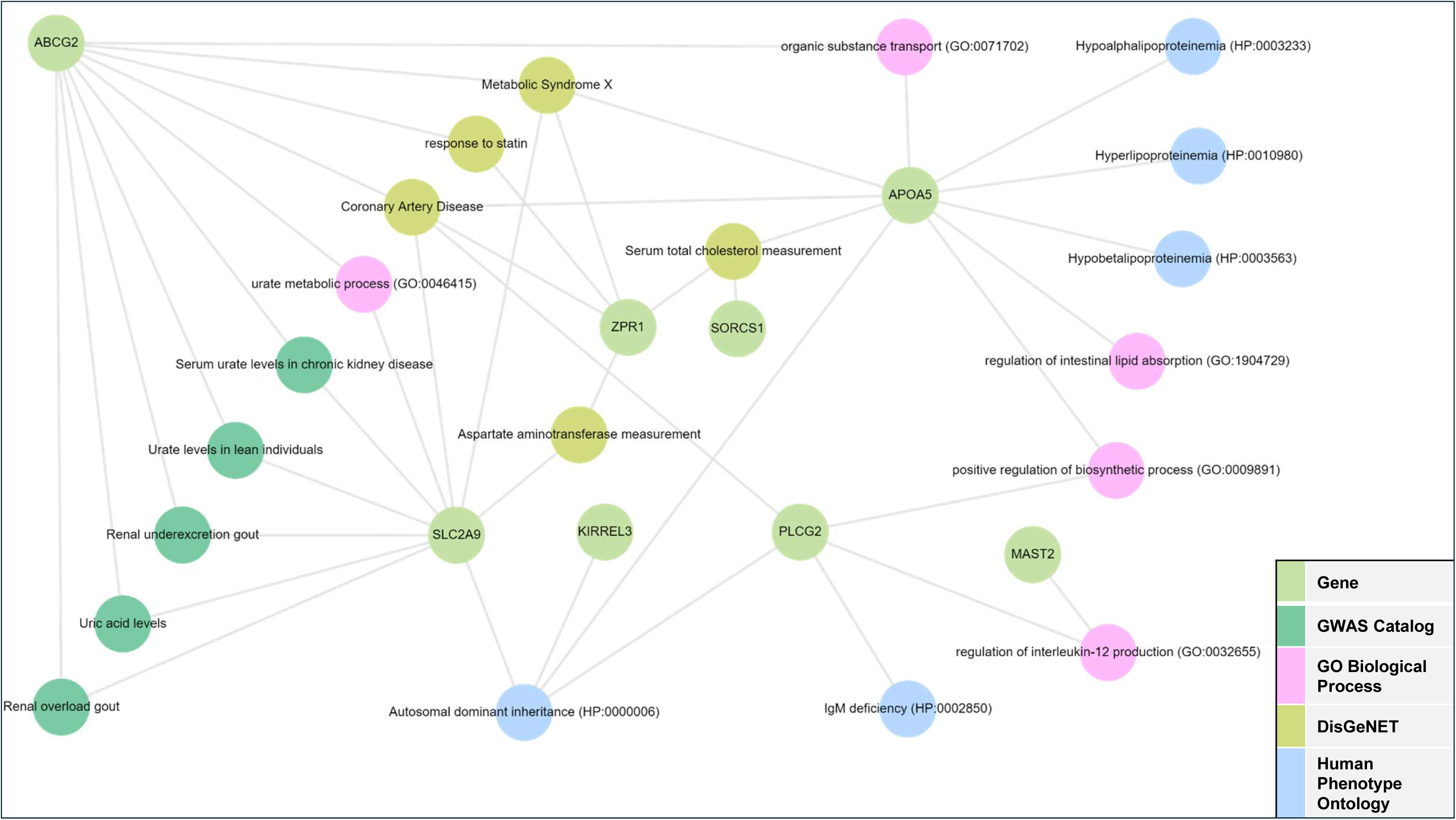
Subnetwork of top genes linked to enriched terms This subnetwork illustrates the top five enriched terms (FDRs < 0.05) for each library, connected to the 11 genes significantly associated with the SGA-induced metabolic side effects, as identified through the primary GWAS and gene-level MAGMA analyses.

#### 3.3.2 Gene set analysis

Fourteen gene sets were nominally associated with SGA-induced metabolic changes. After FDR correction (FDR < 0.05), four gene sets remained significant (**Supplementary Table S6**). The top gene set, *skeletal muscle satellite cell diflerentiation* (p_Bonferroni_ = 1.29e-05, FDR = 4.4e-04), was associated with SGA-induced TG changes^62^. The *mRNA editing* (p_Bonferroni_ = 2.20e-05, FDR = 4.4e-04) gene set was associated with clozapine-induced LDL changes. The gene sets *ER ubiquitin ligase complex* (p_Bonferroni_ = 0.004, FDR = 0.04) and *Saccadic smooth pursuit* (p_Bonferroni_ = 0.004, FDR = 0.04) were associated with clozapine-induced BMI and amisulpride-induced BMI changes respectively.

## 4. Discussion

This study is one of the largest longitudinal PGx GWAS investigations, identifying the genetic variants associated with the lipid and BMI changes induced by seven commonly used SGAs in a Chinese SCZ cohort. Our investigation included 19 316 prescription records and 3 917 to 7 596 metabolic measurements for each outcome, with a median follow-up duration of 5.7 years (SD=3.3, max=18.7), surpassing the duration of comparable GWASs^18, 20–22^. The study design incorporates several key strengths that enhance the robustness and applicability of our findings.

Notably, our cohort recruited from the early psychosis intervention clinic comprised a high proportion of antipsychotic-naïve patients (approximately 63%) at baseline; as such, confounding by previous medications was reduced and likely lower than many other comparable studies, including Adkins *et al*.^18^. Furthermore, our focus on a homogeneous ethnic Chinese sample provides valuable insights specific to this population, which has been underrepresented in previous studies. This is particularly important given the known differences in allele frequency and LD patterns between East Asian (EAS) and European (EUR) populations^63–66^, as evidenced in **Table 2**.

We employed a sophisticated analytical approach using within-subject random effects of SGA-induced lipid/BMI changes. This method substantially reduces the risk of confounding by indication/contraindication^67^, enabling a more accurate assessment of the SGA-induced metabolic side effects. To further mitigate potential confounding effects of other medications, we included lipid-lowering drugs as covariates in the GWAS phenotype estimations. Notably, the mean age of our cohort at the first clinical visit (28.3 years, SD = 9.8) was lower than that reported in a similar GWAS study (mean=40.9 years old, SD=11)^18^, reducing the influence of age-related metabolic changes on our findings.

Our findings might have important clinical implications and contribute to understanding the biological mechanisms underlying SGA-induced metabolic changes. The interindividual variations in PGx may explain the high variability in CVD risk in SCZ patients taking SGAs. Notably, we identified several GW-significant SNPs (rs6532055 in *ABCG2,* rs2644520 near *SORCS1*, rs115843863 near *UPP2*, rs2514895 near *KIRREL3*, and rs1884050603 in *SLC2A9)* and several loci with suggestive evidence (FDR < 0.2). These discoveries may enable more accurate predictions of the risk of metabolic side effects and pave ways for future personalized prescriptions to patients.

The top GW-significant SNP rs6532055 is located in *ABCG2*, which encodes a translocation protein involved in the efflux of antipsychotics across cellular membranes, including the blood‒brain barrier^68, 69^. Its association with olanzapine-induced LDL changes suggests a potential role in antipsychotic pharmacokinetics and lipid metabolism. Notably, *ABCG2* has also been reported to be associated with the degree of LDL reduction in response to rosuvastatin^70, 71^.

Another top gene identified was *SORCS1* which was associated with aripiprazole-induced triglyceride changes. *SORCS1* encodes a member of the VPS10 domain-containing receptor protein family and is strongly expressed in the central nervous system^72^. These receptors bind neuropeptides and facilitate intracellular trafficking. *SORCS1* has been implicated in insulin regulation and type 2 diabetes risk in both animal and clinical studies^73–75^. Notably, several studies have shown that increased TG levels are associated with increased type 2 diabetes risk and impaired fasting glucose^76–78^. Its role in energy balance further supports its potential involvement in antipsychotic-induced metabolic alterations.^79^.

In our additional analyses (**Table 2**), *UPP2* was linked to clozapine-induced HDL changes which encode uridine phosphorylase 2. Notably, several studies have revealed an association between uridine metabolism with lipid metabolism and glucose homeostasis^80–82^. It has been demonstrated that increasing endogenous hepatic uridine levels by inhibiting uridine phosphorylase 2 may reduce drug-induced liver lipid accumulation^82, 83^, although long-term uridine consumption might promote liver lipid accumulation and exacerbate glucose intolerance^82^.

*KIRREL3* encodes a synaptic cell adhesion molecule essential for the formation of target-specific synapses and is expressed in fetal and adult brain tissues. This gene was associated with paliperidone-induced LDL changes in our study and is known primarily for its role in neurological and cognitive disorders, neuroticism, and autism spectrum disorders^84–87^. While its role in lipid metabolism remains to be investigated, this finding suggests a potential novel link between neuronal function and metabolic regulation.

*SLC2A9* encodes glucose transporter 9 (GLUT9), a protein involved in reabsorbing or excreting urate and glucose in kidney proximal tubules. This gene has been strongly associated with uric acid levels and gout in numerous studies^88–91^. SLC2A9 was identified to be linked to TG changes in our sample; notably, studies have revealed a significant positive association between TG and urate levels^92–94^. Recently, a GWAS from Qatar revealed the association of *SLC2A9* with LDL levels^95^.

Additionally, we identified eight SNPs with suggestive evidence (FDR < 0.2) in our primary analyses under an additive model, including SNPs located in or near *APOE*, *MBL2*, *MZT1*, *LOC105373454*, *CDCA7*, *DDX1*, *CD34*, and *SIPA1L2* (Supplementary **Table S2**). Many of these genes are associated with lipid levels, diabetes, CVD, urate levels or other metabolic measurements based on data from the Open Target Platform^45^. These SNPs may also play a role in SGA-induced lipid/BMI changes, but further studies are needed. Similar evidence was found for the 17 suggestive SNPs (FDR < 0.2) identified in our additional analyses using non-additive genetic models (**Supplementary Table S3**). Our MAGMA gene-level analyses identified six GW-significant genes associated with SGA-induced lipid/BMI changes (**Table 4**). Notably, *ABCG2* was identified via both GWAS and MAGMA analyses, providing further support for its potential role in olanzapine-induced LDL changes.

Our study has several limitations. First, only seven SGAs were included, although these are probably among the most commonly prescribed. Future research should aim to expand the scope to include a broader range of SGAs. Second, although our sample sizes (ranging from 43 to 307 participants) are relatively large compared with those of similar GWASs^19–22, 96^, a larger cohort would further increase the statistical power and robustness of our findings. Third, potential residual confounding can also be a problem affecting the estimation of the metabolic side effects of SGAs, which may in turn affect the estimation of the genetic influence on these side effects. While we have applied sophisticated methods such as within-subject random effects estimates and controlled for other concomitant medications, there may be unmeasured confounders that could impact our results. Finally, lifestyle factors such as diet, exercise, alcohol consumption and tobacco smoking were not measured and may be included in future studies. Despite these limitations, our study provides a solid foundation for future research in this critical area of psychopharmacology. Addressing these limitations in subsequent investigations will further refine our understanding of the genetic basis of SGA-induced metabolic side effects.

Our study provides valuable insights into the pharmacogenetics of SGA-induced metabolic changes in a Chinese SCZ cohort. The identified genetic markers not only enhance our understanding of the biological mechanisms underlying these metabolic changes but also hold promise for developing more tailored and safer treatment strategies for individuals with SCZ. However, further studies and replication are needed before these genetic findings can be applied in clinical practice.

## Data Availability

All data produced in the present study are available upon reasonable request to the authors

## Acknowledgments

We thank all collaborators who were involved in patient recruitment and preparation of data, including Ms Hera Yeung, Ms Kirby Tsang, Dr Wong Ting Yat, Dr Karen Ho, Dr Karen Hung, Dr Eric Cheung and Dr KM Cheng. The authors made use of Claude to edit this article for grammatical accuracy, and the original draft was written by the authors without the help of Claude. Claude 3.5 Sonnet was accessed from https://claude.ai and used without modification in June 2024.

## Availability of Data and Materials

The data that support the findings of this study are available upon request from the corresponding authors, Hon-Cheong SO and Simon Sai-Yu LUI. The data are not publicly available because they contain information that could compromise the privacy of research participants.

