## Supplementary Tables and Figures for "Pharmacogenetic Study of Antipsychotic-Induced Lipid and BMI Changes in Chinese Schizophrenia Patients: A Genome-Wide Association Study"

**Table S1 Sample characteristics in our GWAS analyses.**

| Demographics | Cohort | |
|---|---|---|
| | TC,LDL,HDL,TG (N=625) | BMI (N=646) |
| **No. of male** | 290 | 300 |
| **Percentage of male** | 46% | 46% |
| **Mean age at the 1st clinical visit (SD)** | 28.5 (9.6) | 28.3 (9.8) |
| **Mean years of follow-up (SD)** | 5.7 (3.3) | 5.5 (3.2) |

**Table S2 The suggestive SNPs (FDR < 0.2) identified in our primary GWAS analyses.**

| SGA | Phenotype | Test model[1] | SNP | Gene | Nearest | Location | CHR | POS (GRCh38) | Effect Allele | N | Effect AF | 1KG AF (EAS) | 1KG AF (EUR) | Imp. Rsq | Beta | SE | P-value | FDR | $\lambda_{GC}$ (50, 70, 90 percentiles) | Associated diseases (association scores: 0-1) from Open Target Platform |
|---|---|---|---|---|---|---|---|---|---|---|---|---|---|---|---|---|---|---|---|---|
| **Olanzapine** | **LDL** | Additive | rs7412 | *APOE* | *APOE* | Exon (Missense) | 19 | 44908822 | T | 230 | 0.10 | 0.10 | 0.06 | 0.97 | 0.866 | 0.158 | 1.24E-07 | 0.182 | 1.00, 1.00, 1.01 | Hyperlipoproteinemia type 3(0.79), CAD(0.78), Alzheimer disease(0.71) |
| | | | rs2384157 | - | *MBL2* | Intergene | 10 | 53213998 | A | 230 | 0.50 | 0.54 | 0.26 | 0.43 | -0.450 | 0.090 | 1.26E-06 | 0.195 | | Blood protein measurement(0.77), neurodegenerative disease (0.23) |
| | | | rs74625905 | - | *MZT1* | Intergene | 13 | 72402915 | G | 230 | 0.06 | 0.07 | 0.06 | 0.89 | 0.915 | 0.187 | 1.85E-06 | 0.195 | | Cortical surface area measurement (0.35), mathematical ability (0.33) |
| | | | rs56349742 | *LOC105373454* | *RDH14* | Non coding intron | 2 | 18409116 | G | 230 | 0.23 | 0.24 | 0.05 | 0.99 | -0.531 | 0.109 | 1.98E-06 | 0.195 | | Urate measurement(0.62), T2DM(0.51), brain measurement(0.51) |
| **Quetiapine** | **HDL** | Additive | rs2358259 | - | *CDCA7* | Integene | 2 | 173483315 | G | 93 | 0.94 | 0.92 | 0.99 | 0.65 | 1.450 | 0.265 | 5.12E-07 | 0.123 | 0.98, 0.99, 1.00 | ICF syndrome (0.7), mathematical ability(0.59), LDL measurement(0.44) |
| | | | rs10174314 | - | *DDX1* | Integene | 2 | 15752986 | G | 93 | 0.19 | 0.20 | 0.06 | 0.90 | 0.952 | 0.179 | 9.01E-07 | 0.123 | | Urate measurement(0.6), neurodegenerative disease (0.37), Alzheimer disease(0.21) |
| | | | rs117416034 | - | *CD34* | Integene | 1 | 208012181 | A | 93 | 0.02 | 0.02 | 0.00 | 0.97 | 2.920 | 0.551 | 9.98E-07 | 0.123 | | Risk-taking behaviour(0.67), urate measurement(0.58), base metabolic rate(0.41) |
| | | | rs6424242 | *SIPA1L2* | *SIPA1L2* | Upstream gene | 1 | 232632019 | T | 93 | 0.34 | 0.41 | 0.44 | 0.91 | -0.675 | 0.130 | 1.63E-06 | 0.186 | | Pakinson disease(0.66), antisocial behaviour measurement(0.39), ADHD(0.39) |

**Abbreviation:** CHR Chromosome, POS Position, SGA Second generation antipsychotics, SNP Single nucleotide polymorphism, AF Allele frequency, 1KG 1000 Genomes project, "Imp. Rsq" Genotype imputation r-squared, $\lambda_{GC}$ Genomic control inflation factor, CAD Ccoronary artery disease, ADHD Attention deficit hyperactivity disorder.

**Remark:**

[1] Test model indicates the genetic model employed when conducting GWAS using PLINK2. For details, pls refer to the manual of the PLINK2.

**Table S3 Seventeen suggestive SNPs (FDR < 0.2) associated with the SGA-induced lipid levels and BMI changes in our additional analyses.**

| SGA | Trait | Test model[1] | SNP | Gene | Nearest | Location | CHR | POS (GRCh38) | Effect Allele | N | Effect AF | 1KG AF (EAS) | 1KG AF (EUR) | Imp. Rsq | Beta | SE | P-value | FDR | $\lambda_{GC}$ (50, 70, 90 percentiles) | Associated diseases (association scores: 0-1) from Open Target Platform |
|---|---|---|---|---|---|---|---|---|---|---|---|---|---|---|---|---|---|---|---|---|
| **Clozapine** | **TC** | Dominant | rs6495163 | - | *C15orf39* | Intergene | 15 | 75172710 | A | 119 | 0.32 | 0.60 | 0.60 | 0.41 | 0.4070. | 0.162 | 9.69E-08 | 0.184 | 0.98, 0.99, 1.00 | HDL measurement(0.21) |
| | **HDL** | Genotypic | rs1368256 | *LINC00635* | *BBX* | Non coding intron | 3 | 107867577 | T | 119 | 0.87 | 0.84 | 0.51 | 0.55 | NA | NA | 1.48E-07 | 0.089 | 1.01, 1.01, 1.01 | - |
| | | | rs28733194 | *LOC105375377* | *PCLO* | Upstream gene | 7 | 83177233 | T | 119 | 0.68 | 0.60 | 0.34 | 0.80 | NA | NA | 3.19E-07 | 0.089 | | Bipolar disorder(0.64), pontocerebellar hypoplasia type 3(0.62), SCZ(0.56) |
| **Olanzapine** | **TC** | Recessive | rs9634863 | - | *LOC124903233* | Intergene | 13 | 55135136 | G | 230 | 0.72 | 0.75 | 0.53 | 0.89 | -1.369 | 0.282 | 2.31E-06 | 0.097 | 1.00, 1.01, 1.00 | - |
| | | | rs11051856 | *BICD1* | *BICD1* | Intron | 12 | 32217185 | T | 230 | 0.32 | 0.30 | 0.16 | 0.72 | 1.128 | 0.237 | 3.54E-06 | 0.144 | | DNA methylation(0.51), self-reported education(0.35), brain measurement(0.26) |
| | | | rs12548605 | *CSMD1* | *CSMD1* | Intron | 8 | 3649503 | C | 230 | 0.13 | 0.17 | 0.09 | 0.55 | 2.366 | 0.504 | 4.73E-06 | 0.173 | | Body height(0.74), body fat percentage(0.73), estrogen measurement (0.65) |
| | | | rs822205 | - | *UBE2E2* | Intergene | 3 | 23057000 | G | 230 | 0.66 | 0.66 | 0.82 | 0.77 | -0.993 | 0.212 | 4.81E-06 | 0.175 | | T2DM(0.77), HbA1c measurement(0.61), body fat percentage(0.57) |
| **Risperidone** | **TG** | Recessive | rs10911704 | *CBSLR* | *IVNS1ABP* | Non coding intron | 1 | 185322294 | G | 116 | 0.22 | 0.25 | 0.42 | 0.97 | -2.347 | 0.420 | 1.90E-07 | 0.133 | 0.96, 0.98, 0.99 | - |
| | **BMI** | Recessive | rs1494739 | *GADL1* | *GADL1* | Intron | 3 | 30844662 | T | 232 | 0.57 | 0.62 | 0.33 | 0.83 | -0.880 | 0.160 | 9.60E-08 | 0.158 | | Neurofibrillary tangles measurement(0.45), neurodegenerative disease(0.24) |
| **Paliperidone** | **LDL** | Recessive | rs1227097 | *CDH23* | *C10orf105* | Intron | 10 | 71681784 | G | 43 | 0.34 | 0.35 | 0.39 | 0.65 | -2.250 | 0.365 | 8.61E-07 | 0.149 | 0.94, 0.88, 0.97 | Deafness(0.7), Usher syndrome(0.69), antisocial behaviour measurement(0.49) |
| | | | rs7811852 | *AMPH* | *AMPH* | Intron | 7 | 38591056 | C | 43 | 0.55 | 0.57 | 0.50 | 0.48 | -1.990 | 0.344 | 2.53E-06 | 0.160 | | Endocrine system disease(0.53), metabolic disease(0.53), nutritional disorder(0.53) |
| | | Genotypic | rs188559 | *RAB38* | *RAB38* | Intron | 11 | 88160693 | G | 43 | 0.54 | 0.49 | 0.71 | 0.89 | NA | NA | 9.27E-08 | 0.048 | 0.97, 0.99, 1.03 | Calcium measurement(0.72), self-reported education(0.6) |
| | | | rs10256224 | - | *NFE2L3* | Intergene | 7 | 25897720 | T | 43 | 0.24 | 0.22 | 0.20 | 0.60 | NA | NA | 2.90E-07 | 0.049 | | LDL measurement(0.57), TG measurement(0.52), BMI-adjusted waist-hip ratio(0.49) |
| | | | rs10083413 | *CDH23* | *C10orf105* | Intron | 14 | 89464073 | T | 43 | 0.31 | 0.21 | 0.26 | 0.65 | NA | NA | 1.07E-06 | 0.102 | | Deafness(0.7), Usher syndrome(0.69), antisocial behaviour measurement(0.49) |
| | | | rs11146967 | *FOXN3* | *FOXN3* | Intron | 12 | 132605822 | G | 43 | 0.70 | 0.62 | 0.44 | 0.63 | NA | NA | 1.35E-06 | 0.102 | | Glucose measurement(0.78), HDL measurement(0.74), neurodegenerative disease(0.47) |
| | | | rs12410898 | *C1R* | *C1R* | Intron | 1 | 208048939 | A | 43 | 0.15 | 0.11 | 0.02 | 0.82 | NA | NA | 2.79E-06 | 0.179 | | Ehlers-Danlos syndrome(0.65), immunodeficiency(0.55) |
| | | | rs183132876 | *LRCOL1* | *LRCOL1* | Intron | 2 | 66644006 | A | 43 | 0.24 | 0.30 | 0.03 | 0.52 | NA | NA | 4.79E-06 | 0.179 | | TC measurement(0.42), neurodegenerative disease(0.27) |

**Abbreviation:** CHR Chromosome, POS Position, SGA Second generation antipsychotics, SNP Single nucleotide polymorphism, AF Allele frequency, 1KG 1000 Genomes project, "Imp. Rsq" Genotype imputation r-squared, $\lambda_{GC}$ Genomic control inflation factor, T2DM Type 2 diabetes mellitus.

**Remark:**

[1] Test model indicates the genetic model employed when conducting GWAS using PLINK2. For details, pls refer to the manual of the PLINK2.

**Table S4 GW-significant and suggestive SNPs associated with the SGA-induced lipid levels and BMI changes in additional analyses (heterozygous model).**

| **Test model[1]** | **Heterozygous only** | | | | | | | |
|---|---|---|---|---|---|---|---|---|
| **SGA** | **Clozapine** | **Aripiprazole** | **Olanzapine** | | | | | |
| **Phenotype** | **HDL** | **TC** | **BMI** | | | | | |
| **SNP** | rs115843863 | rs34704719 | rs2021526 | rs9343212 | rs743506 | rs7634074 | rs4880783 | rs76896475 |
| **Gene** | - | *PLCG2* | *ENC1* | *LOC105377858* | *NOS3* | *PCOLCE2* | *ADARB2* | - |
| **Nearest** | *UPP2* | - | *HEXB* | *COL12A1* | *ATG9B* | *PCOLCE2* | *IDI1* | *TMEM87B* |
| **Location** | Intergene | Intron | Intron | Non coding intron | Intron | Intron | 3 prime UTR | Intergene |
| **CHR** | 2 | 16 | 5 | 6 | 7 | 3 | 10 | 2 |
| **POS (GRCh38)** | 157988744 | 81794411 | 74637753 | 74724956 | 151009827 | 142849084 | 1177802 | 112036511 |
| **Effect Allele** | T | C | C | C | A | T | T | T |
| **N** | 119 | 238 | 307 | 307 | 307 | 307 | 307 | 307 |
| **Effect AF** | 0.19 | 0.66 | 0.73 | 0.76 | 0.88 | 0.35 | 0.59 | 0.02 |
| **1KG AF (EAS)** | 0.20 | 0.66 | 0.67 | 0.74 | 0.80 | 0.33 | 0.64 | 0.04 |
| **1KG AF (EUR)** | 0.03 | 0.19 | 0.70 | 0.62 | 0.71 | 0.70 | 0.79 | 0.00 |
| **Imp. Rsq** | 0.90 | 0.49 | 0.41 | 0.65 | 0.51 | 0.93 | 0.75 | 0.47 |
| **Beta** | 1.102 | 0.734 | -0.599 | -0.609 | -0.639 | 0.545 | 0.536 | -1.328 |
| **SE** | 0.171 | 0.127 | 0.115 | 0.120 | 0.132 | 0.113 | 0.112 | 0.279 |
| **P-value** | **3.52E-09** | **2.30E-08** | 3.38E-07 | 6.99E-07 | 2.00E-06 | 2.44E-06 | 2.83E-06 | 3.06E-06 |
| **FDR** | 0.008 | 0.161 | 0.124 | 0.124 | 0.167 | 0.167 | 0.167 | 0.169 |
| **$\lambda_{GC}$ (50, 70, 90 percentiles)** | 1.02, 1.01, 1.00 | 1.00, 1.01, 1.01 | 1.01, 1.00, 1.00 | | | | | |
| **QQPlot** | 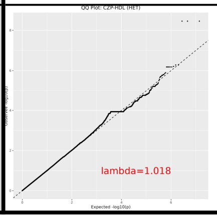  | 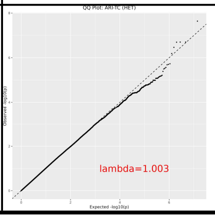  | 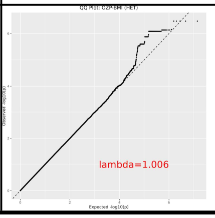  | | | | | |

**Abbreviation:** CHR Chromosome, POS Position, SGA Second generation antipsychotics, SNP Single nucleotide polymorphism, AF allele frequency, 1KG 1000 Genomes project, "Imp. Rsq" Genotype imputation r-squared, $\lambda_{GC}$ Genomic Control Inflation Factor

**Remark:**

[1] Test model indicates the genetic model employed when conducting GWAS using PLINK2. For details, pls refer to the manual of PLINK2.

**Table S5 P-value and FDR of the significant terms identified in gene set enrichment analyses via the Enrichr-KG platform.**

| Term | Library | p-value | FDR |
|---|---|---|---|
| Renal overload gout | GWAS_Catalog_2019 | 2.7E-06 | 2.8E-04 |
| Serum urate levels in chronic kidney disease | GWAS_Catalog_2019 | 5.8E-06 | 2.9E-04 |
| Renal underexcretion gout | GWAS_Catalog_2019 | 9.9E-06 | 3.4E-04 |
| urate metabolic process (GO:0046415) | GO_Biological_Process_2021 | 1.5E-05 | 3.4E-03 |
| response to statin | DisGeNET | 6.9E-05 | 1.5E-02 |
| Coronary Artery Disease | DisGeNET | 1.0E-04 | 1.5E-02 |
| Aspartate aminotransferase measurement | DisGeNET | 1.1E-04 | 1.5E-02 |
| Uric acid levels | GWAS_Catalog_2019 | 1.3E-04 | 3.2E-03 |
| Metabolic Syndrome X | DisGeNET | 1.6E-04 | 1.6E-02 |
| Urate levels in lean individuals | GWAS_Catalog_2019 | 1.9E-04 | 3.4E-03 |
| positive regulation of biosynthetic process (GO:0009891) | GO_Biological_Process_2021 | 2.6E-04 | 2.6E-02 |
| regulation of interleukin-12 production (GO:0032655) | GO_Biological_Process_2021 | 3.5E-04 | 2.6E-02 |
| Serum total cholesterol measurement | DisGeNET | 5.4E-04 | 3.1E-02 |
| organic substance transport (GO:0071702) | GO_Biological_Process_2021 | 2.4E-03 | 3.8E-02 |
| Autosomal dominant inheritance (HP:0000006) | Human_Phenotype_Ontology | 2.5E-03 | 1.8E-02 |
| regulation of intestinal lipid absorption (GO:1904729) | GO_Biological_Process_2021 | 2.7E-03 | 3.8E-02 |
| Hypobetalipoproteinemia (HP:0003563) | Human_Phenotype_Ontology | 4.4E-03 | 1.8E-02 |
| Hyperlipoproteinemia (HP:0010980) | Human_Phenotype_Ontology | 4.9E-03 | 1.8E-02 |
| IgM deficiency (HP:0002850) | Human_Phenotype_Ontology | 5.5E-03 | 1.8E-02 |
| Hypoalphalipoproteinemia (HP:0003233) | Human_Phenotype_Ontology | 5.5E-03 | 1.8E-02 |

**Table S6 Gene sets significantly associated ($p_{bonferroni<}0.05$) with SGA-induced lipid level and BMI changes using MAGMA gene set analyses.**

| SGA | Trait | MSigDB Gene Set | Name of Gene set | $P_{bonferroni}$ [1] | FDR[2,3] |
|---|---|---|---|---|---|
| **Any SGA use** | **TG** | Curated (C5) | GOBP_SKELETAL_MUSCLE_SATELLITE_CELL_DIFFERENTIATION* | 1.29E-05 | **4.40E-04** |
| **Clozapine** | **LDL** | Curated (C2) | REACTOME_MRNA_EDITING* | 2.20E-05 | **4.40E-04** |
| **Clozapine** | **BMI** | Curated (C5) | GOCC_ER_UBIQUITIN_LIGASE_COMPLEX* | 0.004 | **0.04** |
| **Amisulpride** | **BMI** | Curated (C5) | HP_SACCADIC_SMOOTH_PURSUIT* | 0.004 | **0.04** |
| **Quetiapine** | **HDL** | Curated (C5) | GOBP_PROTEIN_IMPORT | 0.006 | 0.09 |
| **Clozapine** | **TC** | Curated (C2) | BIOCARTA_VITCB_PATHWAY | 0.013 | 0.09 |
| **Any SGA use** | **LDL** | Hallmark (H) | HALLMARK_MYC_TARGETS_V1 | 0.013 | 0.07 |
| **Clozapine** | **TC** | Curated (C5) | GOBP_SPONTANEOUS_NEUROTRANSMITTER_SECRETION | 0.019 | 0.09 |
| **Risperidone** | **LDL** | Curated (C2) | REACTOME_DEADENYLATION_OF_MRNA | 0.021 | 0.09 |
| **Clozapine** | **BMI** | Curated (C2) | REACTOME_FATTY_ACIDS | 0.023 | 0.09 |
| **Quetiapine** | **TC** | Hallmark (H) | HALLMARK_WNT_BETA_CATENIN_SIGNALING | 0.024 | 0.09 |
| **Quetiapine** | **BMI** | Curated (C5) | GOBP_TAURINE_METABOLIC_PROCESS | 0.029 | 0.09 |
| **Amisulpride** | **HDL** | Curated (C5) | GOBP_POSITIVE_REGULATION_OF_MYELOID_CELL_DIFFERENTIATION | 0.030 | 0.09 |
| **Risperidone** | **TC** | Curated (C2) | UDAYAKUMAR_MED1_TARGETS_UP | 0.038 | 0.11 |

**Remarks:**

[1] $P_{bonferroni}$ is the adjusted P-value for each gene set, where the number of tests conducted per gene set equals the size of the corresponding MSigDB.

[2] FDR is the FDR-adjusted p-value (based on $p_{bonferroni}$) to control the false discovery rate due to multiple tests (40 in our study) issues.

[3] Only the top four gene sets marked with an asterisk (*) survived a statistically significant threshold (FDR < 0.05).

**Figure S1 The distribution of GWAS phenotypes showing the existence of strong outliers.**

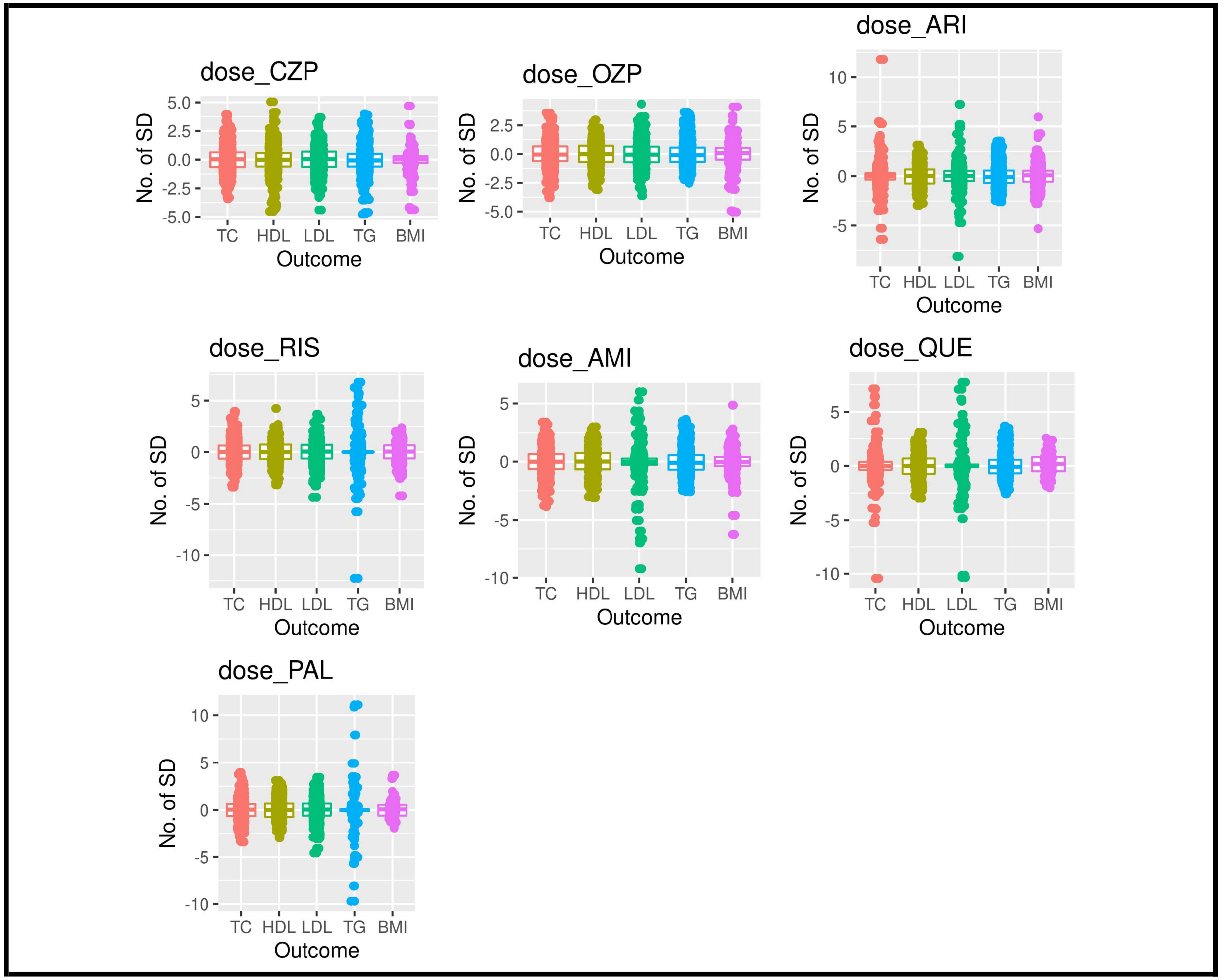

**Figure S2 QQ plots of GWAS with suggestive SNPs achieving FDR < 0.2.**

<table>
<tr><th>Analysis type</th><th>SGA</th><th>Trait</th><th>Genetic model</th><th>QQ Plot</th><th>$\lambda_{GC}$ (50, 70, 90 percentiles)</th></tr>
<tr><td rowspan="3">Primary Analyses</td><td rowspan="2">Olanzapine</td><td>TC</td><td>Additive</td><td>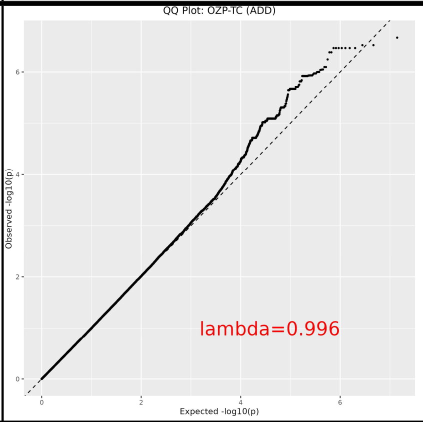
</td><td>1.00, 1.00, 1.01</td></tr>
<tr><td>LDL</td><td>Additive</td><td>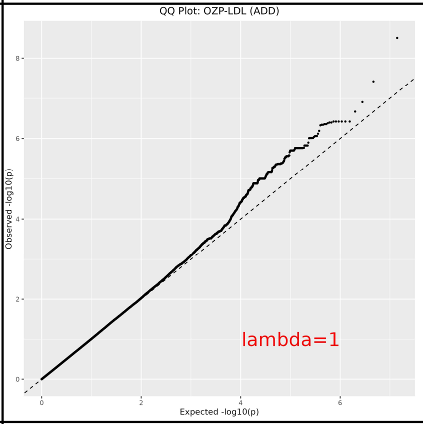
</td><td>1.00, 1.00, 1.01</td></tr>
<tr><td>Quetiapine</td><td>HDL</td><td>Additive</td><td>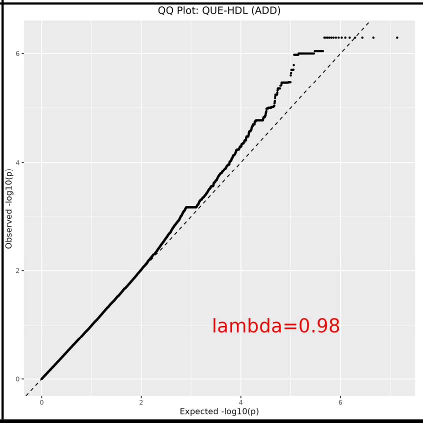
</td><td>0.98, 0.99, 1.00</td></tr>
<tr><td></td><td>Clozapine</td><td>TC</td><td>Dominant</td><td>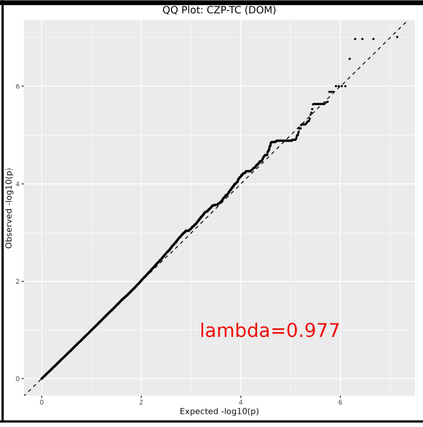
</td><td>0.98, 0.99, 1.00</td></tr>
</table>

| | | | | | |
|---|---|---|---|---|---|
| **Additional Analyses** | **Clozapine** | **HDL** | Genotypic | 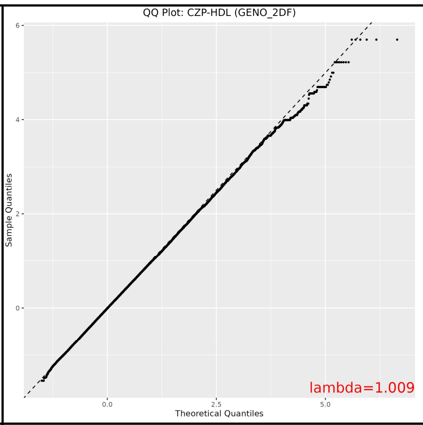 | 1.01, 1.01, 1.01 |
| | **Olanzapine** | **TC** | Recessive | 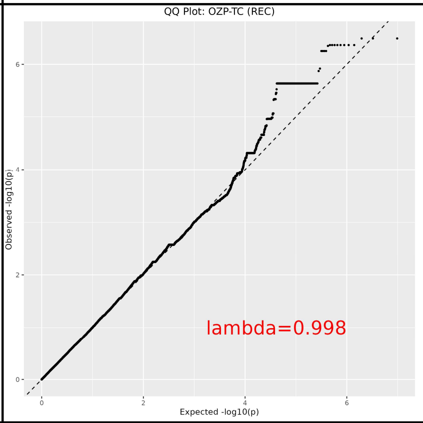 | 1.00, 1.01, 1.00 |
| | **Risperidone** | **TG** | Recessive | 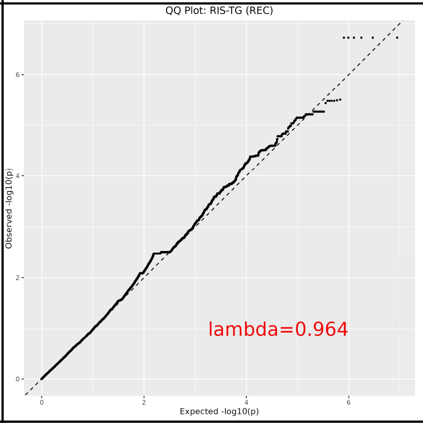 | 0.96, 0.98, 0.99 |
| | | **BMI** | Recessive | 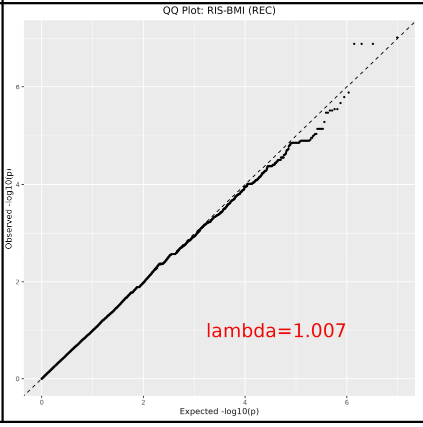 | 1.01, 1.01, 0.99 |

| | | | | | |
|---|---|---|---|---|---|
| | **Quetiapine** | **TG** | Dominant | 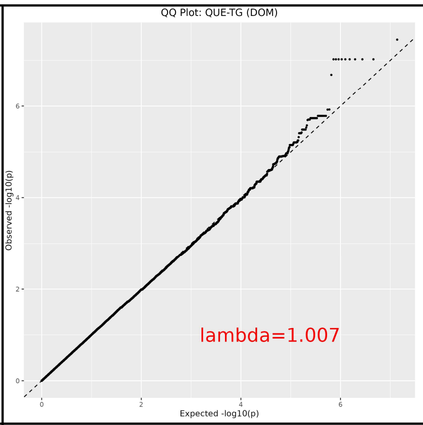 | 1.01, 1.01, 1.01 |
| | **Paliperidone** | **LDL** | Genotypic | 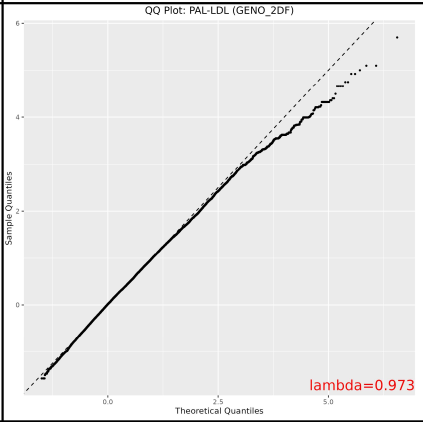 | 0.97, 0.99, 1.03 |
| | | | Recessive | 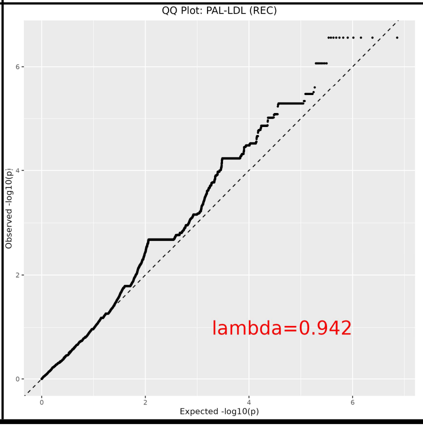 | 0.94, 0.88, 0.97 |

**Figure S3 QQ plots of MAGMA gene-level analyses with GW-significant genes.**

<table>
<tr><th>SGA</th><th>Trait</th><th>QQ Plot</th><th>$\lambda_{GC}$</th></tr>
<tr><td rowspan="2">Olanzapine</td><td>HDL</td><td>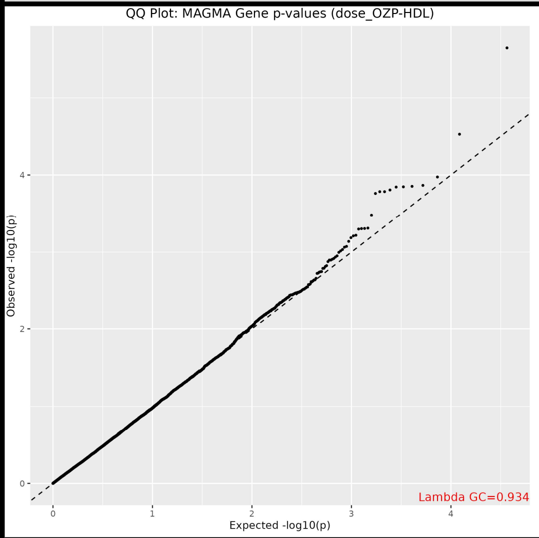
</td><td>0.93</td></tr>
<tr><td>LDL</td><td>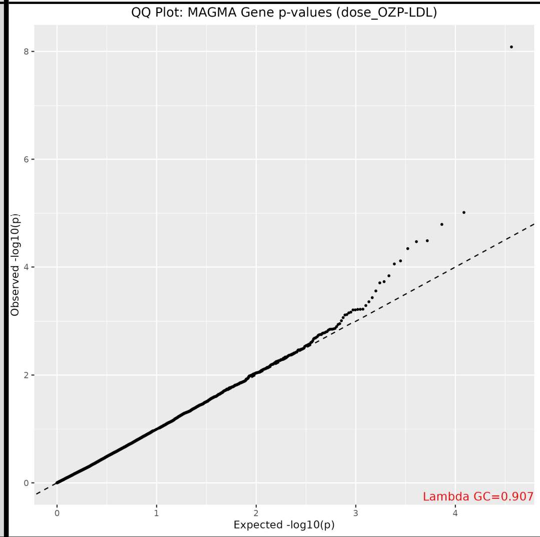
</td><td>0.9</td></tr>
<tr><td>Risperidone</td><td>LDL</td><td>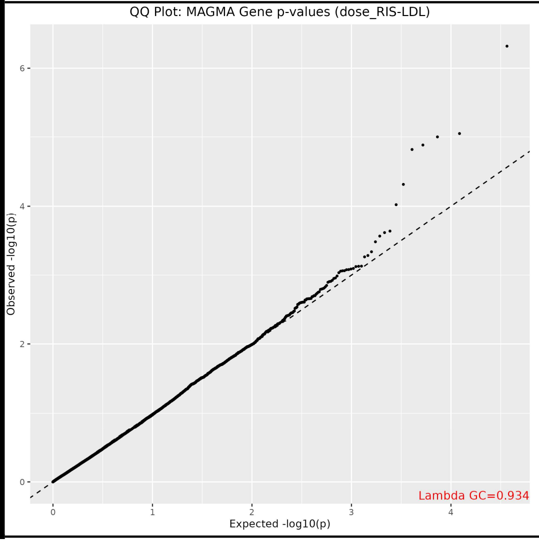
</td><td>0.93</td></tr>
</table>

| | | | |
|---|---|---|---|
| **Quetiapine** | **TG** | 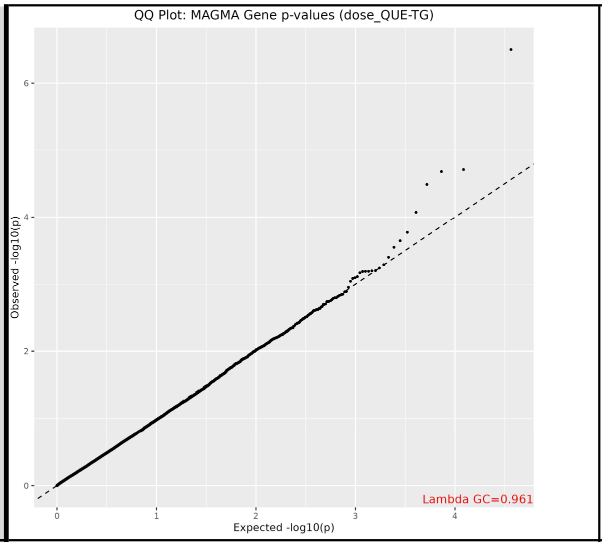 | 0.96 |
| **Any SGA use** | **TG** | 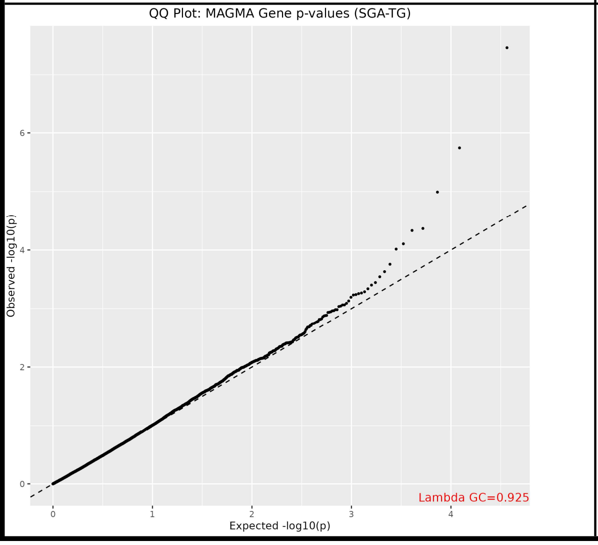 | 0.93 |
