## Supplementary Text for "Pharmacogenetic Study of Antipsychotic-Induced Lipid and BMI Changes in Chinese Schizophrenia Patients: A Genome-Wide Association Study"

### Supplementary Text 1:

### The full formula of 40 random-effect LLM models is expressed in lmer4 (R package) format.

**Random-effect LMM model for CLOZAPINE (one model per outcome)**

Log(TC/HDL/LDL/TG/BMI measure) ~ age + gender + education + tx_duration +

dose.OLANZAPINE + dose.ARIPIPRAZOLE + dose.RISPERIDONE + dose.AMISULPRIDE + dose.QUETIAPINE + dose.PALIPERIDONE +

dose.HALOPERIDOL + dose.TRIFLUOPERAZINE + dose.VALPROATE + dose.LITHIUM + dose.CITALOPRAM + dose.SERTRALINE +

dose.METFORMIN + dose.ATORVASTATIN + dose.SIMVASTATIN +

**bz_dose.CLOZAPINE + wz_dose.CLOZAPINE + (1 + wz_dose.CLOZAPINE | ID)**

**Random-effect LMM model for OLANZAPINE (one model per outcome)**

Log(TC/HDL/LDL/TG/BMI measure) ~ age + gender + education + tx_duration +

dose.CLOZAPINE + dose.ARIPIPRAZOLE + dose.RISPERIDONE + dose.AMISULPRIDE + dose.QUETIAPINE + dose.PALIPERIDONE +

dose.HALOPERIDOL + dose.TRIFLUOPERAZINE + dose.VALPROATE + dose.LITHIUM + dose.CITALOPRAM + dose.SERTRALINE +

dose.METFORMIN + dose.ATORVASTATIN + dose.SIMVASTATIN +

**bz_dose.OLANZAPINE + wz_dose.OLANZAPINE + (1 + wz_dose.OLANZAPINE | ID)**

**Random-effect LMM model for ARIPIPRAZOLE (one model per outcome)**

Log(TC/HDL/LDL/TG/BMI measure) ~ age + gender + education + tx_duration +

dose.CLOZAPINE + dose.OLANZAPINE + dose.RISPERIDONE + dose.AMISULPRIDE + dose.QUETIAPINE + dose.PALIPERIDONE +

dose.HALOPERIDOL + dose.TRIFLUOPERAZINE + dose.VALPROATE + dose.LITHIUM + dose.CITALOPRAM + dose.SERTRALINE +

dose.METFORMIN + dose.ATORVASTATIN + dose.SIMVASTATIN +

**bz_dose.ARIPIPRAZOLE + wz_dose.ARIPIPRAZOLE + (1 + wz_dose.ARIPIPRAZOLE | ID)**

**Random-effect LMM model for RISPERIDONE (one model per outcome)**

Log(TC/HDL/LDL/TG/BMI measure) ~ age + gender + education + tx_duration +

dose.CLOZAPINE + dose.OLANZAPINE + dose.ARIPIPRAZOLE + dose.AMISULPRIDE + dose.QUETIAPINE + dose.PALIPERIDONE +

dose.HALOPERIDOL + dose.TRIFLUOPERAZINE + dose.VALPROATE + dose.LITHIUM + dose.CITALOPRAM + dose.SERTRALINE +

dose.METFORMIN + dose.ATORVASTATIN + dose.SIMVASTATIN +

**bz_dose.RISPERIDONE + wz_dose.RISPERIDONE + (1 + wz_dose.RISPERIDONE | ID)**

**Random-effect LMM model for AMISULPRIDE (one model per outcome)**

Log(TC/HDL/LDL/TG/BMI measure) ~ age + gender + education + tx_duration +

dose.CLOZAPINE + dose.OLANZAPINE + dose.ARIPIPRAZOLE + dose.RISPERIDONE + dose.QUETIAPINE + dose.PALIPERIDONE +

dose.HALOPERIDOL + dose.TRIFLUOPERAZINE + dose.VALPROATE + dose.LITHIUM + dose.CITALOPRAM + dose.SERTRALINE +

dose.METFORMIN + dose.ATORVASTATIN + dose.SIMVASTATIN +

**bz_dose.AMISULPRIDE + wz_dose.AMISULPRIDE + (1 + wz_dose.AMISULPRIDE | ID)**

**Random-effect LMM model for QUETIAPINE (one model per outcome)**

Log(TC/HDL/LDL/TG/BMI measure) ~ age + gender + education + tx_duration +

dose.CLOZAPINE + dose.OLANZAPINE + dose.ARIPIPRAZOLE + dose.RISPERIDONE + dose.AMISULPRIDE + dose.PALIPERIDONE +

dose.HALOPERIDOL + dose.TRIFLUOPERAZINE + dose.VALPROATE + dose.LITHIUM + dose.CITALOPRAM + dose.SERTRALINE +

dose.METFORMIN + dose.ATORVASTATIN + dose.SIMVASTATIN +

**bz_dose.QUETIAPINE + wz_dose.QUETIAPINE + (1 + wz_dose.QUETIAPINE | ID)**

**Random-effect LMM model for PALIPERIDONE (one model per outcome)**

Log(TC/HDL/LDL/TG/BMI measure) ~ age + gender + education + tx_duration +

dose.CLOZAPINE + dose.OLANZAPINE + dose.ARIPIPRAZOLE + dose.RISPERIDONE + dose.AMISULPRIDE + dose.QUETIAPINE +

dose.HALOPERIDOL + dose.TRIFLUOPERAZINE + dose.VALPROATE + dose.LITHIUM + dose.CITALOPRAM + dose.SERTRALINE +

dose.METFORMIN + dose.ATORVASTATIN + dose.SIMVASTATIN +

**bz_dose.PALIPERIDONE + wz_dose.PALIPERIDONE + (1 + wz_dose.PALIPERIDONE | ID)**

The dose of the drug (*dose.<drug>*) represents the time-varying covariate of second-generation antipsychotics (SGAs), first-generation antipsychotics (FGA), and other psychotropic or concomitant drug daily dose (mg) that may affect the lipid/BMI measure. The between-subject and within-subject target SGA doses (mg) are denoted by *bz_dose.<target SGA>* and *wz_dose.<target SGA>,* respectively. These dose-related covariates were recorded 21 days before the lipid or BMI measurements.

The subject identifier (*ID*) serves as the grouping factor, denoted by (……|ID). This indicates that the model is a random-effect (also known as random-slope) linear mixed-effects model (LMM). The random-effect coefficients of *wz_dose.<target SGA>* will be extracted from the fitted model, representing the phenotype in our GWAS and MAGMA analyses.

| **Random-effect LMM model for SGAs - binary variable (one model per outcome)** |
|---|
| Log(TC/HDL/LDL/TG/BMI measure) ~ age + gender + education + tx_duration + |
| FGA + VALPROATE + LITHIUM + CITALOPRAM + SERTRALINE + |
| METFORMIN + ATORVASTATIN + SIMVASTATIN + |
| **bz_SGA + wz_SGA + (1 + wz_SGA \| ID)** |

The binary variable for the drug *(<drug>*) represents the time-varying binary covariate of SGAs, first-generation antipsychotics (FGA), and other psychotropic or concomitant drug daily prescription that may affect the lipid/BMI measure (0 – not prescribed, 1 - prescribed). The between-subject and within-subject SGA binary variables are denoted by *bz_SGA* and *wz_SGA*, respectively. These binary covariates were recorded 21 days before the lipid or BMI measurements.

The subject identifier (*ID*) serves as the grouping factor, denoted by (……|ID). This indicates that the model is a random-effect (also known as random-slope) LMM. The random-effect coefficients of *wz_SGA* will be extracted from the fitted model, representing the phenotype in our GWAS and MAGMA analyses.

### Supplementary Text 2:

### Software and tools used in this project.

| Procedures | Tools |
|---|---|
| Quality control of genotype | PLINK v1.9p |
| Liftover from GRCh37 to GRCh38 | CrossMap v0.6.4 |
| Genotype imputation | Eagle2,<br>Minimac4,<br>ChinaMAP phase1.v1 reference panel |
| Random-effect modelling | R script run on Rstudio v1.4.1106 and R v4.05 with the following R packages:<br>• data.table v1.14.0<br>• dplyr v1.0.6<br>• lme4 v1.1.27<br>• lubridate v1.7.10<br>• zoo v1.8.9 |
| GWAS analyses | PLINK v2.00a |
| Fine mapping | SusieR v0.12.41 |
| Gene-based and gene-set analyses | MAGMA v1.10 |
| Post-GWAS annotation | VEP v111.0 with VEP cache v111_GRCh38. |
| | Open Target Platform |
| | GWAS Catalogue |
| FDR estimation | R package: qvalue v2.15.0 |
